# SARS-CoV-2 virus in Raw Wastewater from Student Residence Halls with concomitant 16S rRNA Bacterial Community Structure changes

**DOI:** 10.1101/2024.02.11.24302582

**Authors:** Y. Li, K. T. Ash, D. C. Joyner, D. E. Williams, T. C. Hazen

**Author notes:** **Correspondence:** Terry C. Hazen. **Author Contributions:** Writing: YL, TCH; Collection: DEW, DCJ; Lab analysis: YL, KTA, DEW, DCJ; Data Analysis: YL, KTA, TCH; Project Management: TCH, DCJ.

## Abstract

The detection of severe acute respiratory syndrome coronavirus 2 (SARS-CoV-2) RNA in sewage is well-established, but the concomitant changes in microbial compositions during the pandemic remain insufficiently explored. This study investigates the impact of the SARS-CoV-2 virus on microbial compositions in raw sewage, utilizing 16S rRNA sequencing to analyze wastewater samples collected from six dormitories over a one-year field trial at the University of Tennessee, Knoxville. The concentration of SARS-CoV-2 RNA was assessed using a reverse transcription-quantitative polymerase chain reaction. Significant variations in bacterial composition were evident across the six dormitories, highlighting the importance of independently considering spatial differences when evaluating the raw wastewater microbiome. Positive samples for SARS-CoV-2 exhibited a prominent representation of exclusive species across all dormitories, coupled with significantly reduced bacterial diversity compared to negative samples. The correlation observed between the relative abundance of enteric pathogens and potential pathogens at sampling sites introduces a significant dimension to our understanding of COVID-19, especially the notable correlation observed in positive SARS-CoV-2 samples. Furthermore, the significant correlation in the relative abundance of potential pathogens between positive and negative SARS-CoV-2 raw sewage samples may be linked to the enduring effects of microbial dysbiosis observed during COVID-19 recovery. These findings provide valuable insights into the microbial dynamics in raw sewage during the COVID-19 pandemic.

## Introduction

The interconnection between sewage and the human gut microbiota has garnered significant interest, revealing a substantial overlap in microbial composition. Newton et al. (2015) noted that the microbial composition of sewage, primarily originating from the human gut, comprises a diverse array of both beneficial and pathogenic species, with bacteria and viruses playing central roles. Robust evidence consistently supports the notable similarity between the microbial profiles of raw sewage and the human gut. Cai et al. (2014) emphasized that the total abundance of high-level genera in influent sewage is nearly 50%, similar to that of the human gut, thus highlighting the human gut as the primary source of bacterial collection in sewage. Newton et al. (2015) further reported that sequences representing approximately 78% of a stool sample comprised around 12% of a sewage sample. Extrapolating this ratio to 100%, their estimation suggests that only 15% of amplicons in a typical sewage sample originate from human stool. However, Fierer et al. (2022) found that bacteria derived from fecal material constitute a relatively small fraction of the taxa in collected samples, underscoring the significance of environmental sources in shaping the sewage microbiome. It is still unclear if raw sewage truly reflects the microbial composition of the human gut.

Crucially, numerous studies have illustrated that respiratory infections associated with COVID- 19 correlate with changes in the composition of the gut microbiota (Gu et al. 2020, Zuo et al. 2020). The dysbiosis of COVID-19 may enhance gut permeability, leading to secondary infections and organ failure. Simultaneously, disruptions in gut barrier integrity could potentially facilitate the translocation of SARS-CoV-2 from the lungs to the intestinal lumen (AKTAŞ and Aslim 2020). Gu et al. (2020) and Zuo et al. (2020) observed that, compared to fecal samples from healthy people, fecal samples from COVID-19 patients had significantly reduced bacterial diversity, a significantly higher relative abundance of opportunistic pathogens and a lower relative abundance of beneficial symbionts. Liu et al. (2022) even found that gut dysbiosis persisted even after clearance of SARS-CoV-2 at 6 months. Patients with COVID-19 exhibit significant alterations in fecal microbiomes, suggesting potential changes in the wastewater microbiome during the pandemic. Currently, research on microbial compositions in wastewater with positive and negative SARS-CoV-2 samples remains limited, with Gallardo-Escárate et al. (2021) being the sole study to explore such dynamics across three sampling communities using nanopore technology. Their findings highlighted a robust association between the microbiota of positive SARS-CoV-2 wastewater samples and enteric bacteria. Notably, integrating the Wastewater-Based Epidemiology tool with metagenomic analysis, employing 16S rRNA sequencing technology to investigate changes in sewage microbiota during the COVID-19 pandemic, remains an unexplored avenue that warrants further research.

This study employs 16S rRNA sequencing to thoroughly analyze microbial compositions in raw sewage samples, differentiating between those with positive and negative COVID-19 status. The primary goal is to identify distinct patterns or shifts in the bacterial community associated with the presence of the virus. Through the utilization of this technology, the research aims to provide a nuanced understanding of the dynamics of viral shedding, microbial interactions, and the overall impact of SARS-CoV-2 on the sewage microbiome over a year-long field trial conducted in six campus dormitories. Including COVID-19-negative sewage samples as a control allows for identifying specific changes attributable to viral presence, facilitating the establishment of correlations between the sewage microbiota and COVID-19 prevalence in human communities. Essentially, this investigation seeks to address the knowledge gap regarding the interplay between SARS-CoV-2 and the sewage microbiome, offering valuable insights into the potential utility of wastewater-based epidemiology for monitoring and assessing COVID-19 prevalence.

## Materials and Methods

### Raw Sewage Sampling and Sample Processing

Raw wastewater was systematically collected from six student residence halls on the University of Tennessee, Knoxville campus, as illustrated in Figure 1. Each of these residential dormitories accommodated a population of over 400 students, and a detailed summary of their characteristics is presented in Table 1. Sampling was from access points to the main sewage pipe in the basement of the building or at the first access point to a raw sewer manhole immediately outside the building, specifically before the convergence or merging with other sewer conduits. This sampling initiative occurred from September 14, 2020, to October 11, 2021.

**Figure 1.**
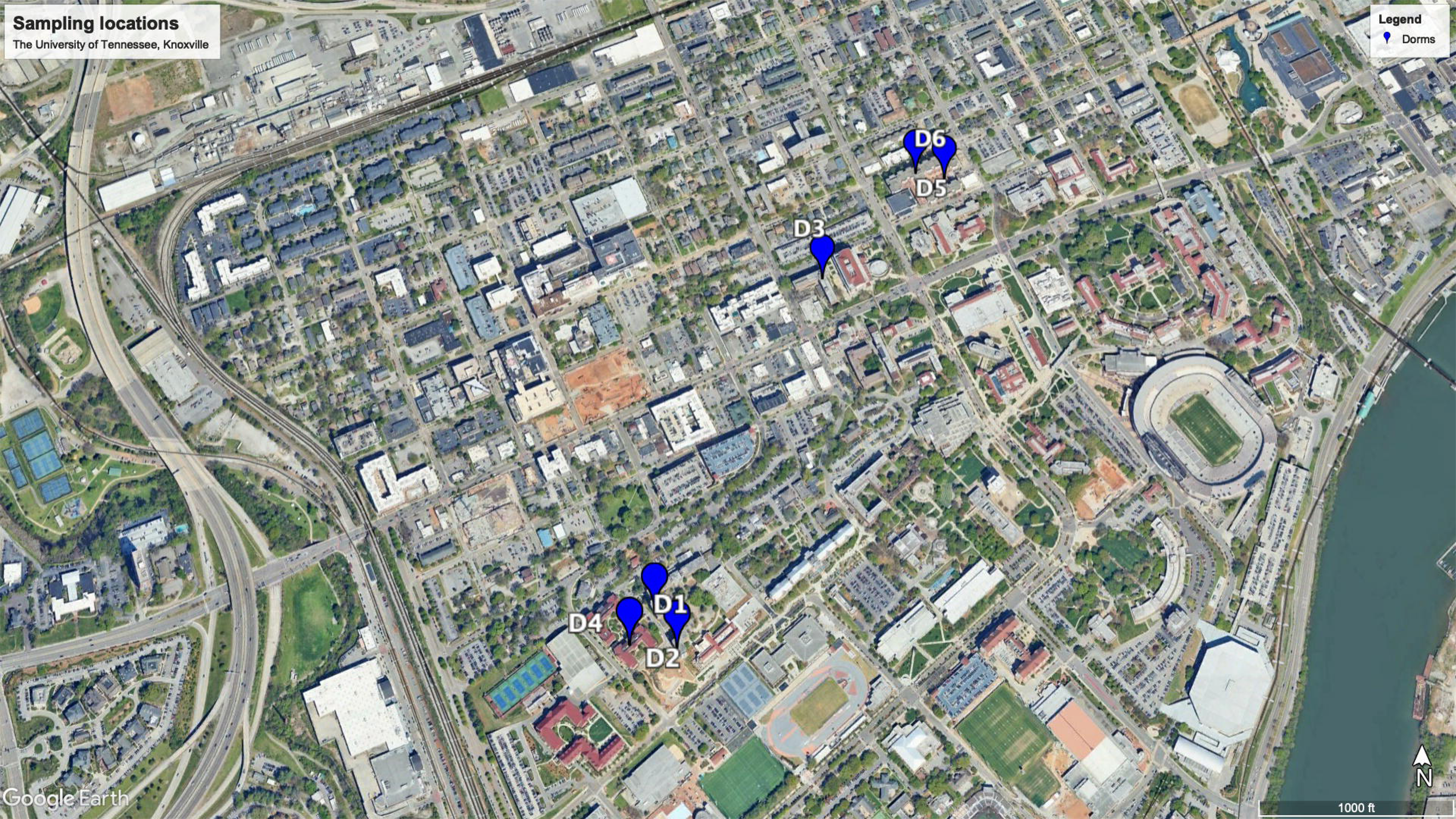
Map of the sampling locations on the University of Tennessee-Knoxville campus.

**Table 1.** Demography data for D1, D2, D3, D4, D5, and D6.

| Sampling site | Sampling Point | Gender | Student Number |
| --- | --- | --- | --- |
| D1 | Direct Dispense from the valve | Male | 387-504 |
| D2 | Direct Dispense from the valve | Female | 469-531 |
| D3 | Direct Dispense from the valve | Mix | 254-279 |
| D4 | Direct Dispense from the valve | Mix | 529-637 |
| D5 | Direct Dispense from the valve | Mix | 10-672 |
| D6 | Direct Dispense from the valve | Mix | 580-672 |

Grab samples (>50 ml) were collected at the manhole using a stainless-steel telescopic rod pole swivel dipper water swing sampler. Alternatively, samples were obtained from the valve by submerging a sterile Nalgene bottle into the flowing sewage. Sampling commenced at 8:00 am, and all collected samples were promptly transported to the BSL-2 laboratory in a cooler with ice. The transit time was kept to less than 3 h to ensure immediate processing.

Upon reaching the laboratory, sewage samples underwent pasteurization for 2 h at 60°C. Following pasteurization, centrifugation at 5,000 x g for 10 min occurred, and subsequent filtration was carried out through sequentially sized 0.45 µm and 0.22 µm nitrocellulose filters. These filters were individually placed in DNA LoBind tubes and stored at -80°C until DNA extraction. Concentration was achieved using an Amicon Ultra-15 filtration device, with centrifugation at either 4,000 x g for 30 min (Swing-arm rotor) or 5,000 x g for 20 min (Fixed-angle rotor) at room temperature. The resulting concentrated solution, approximately 250 μL, was carefully transferred to 2 mL DNA LoBind tubes.

RNA extraction was performed using the Qiagen viral RNA Mini Kit, following the instructions of the manufacturer, yielding 60 μL of extracted RNA, with a negative control using DNase/RNase-free water. Subsequently, the RNA samples were stored at -80°C and subjected to RT-qPCR analysis within 24 h following extraction (Ash et al. 2023, Li et al. 2023).

### RT-qPCR

To quantify the concentrations of SARS-CoV-2 and PMMoV RNA in each sample, we employed RT-qPCR. Specifically, we measured SARS-CoV-2 N1 using the TaqPath 1-Step RT-qPCR Master Mix, CG (Thermo Fisher Scientific) on an Applied Biosystems QuantStudios 7 Pro Real-Time PCR System instrument. Each 20 μL reaction mixture comprised 5 μL of 4X Master Mix (Thermo Fisher Scientific), 0.25 μL of a 10 μmol/L probe, 1 μL each of 10 μmol/L forward and reverse primers, 7.75 μL of nuclease-free water, and 5 μL of nucleic acid extract. After accurate pipetting of reagents into 96-well plates, a 10-second vortex mixing step followed. The RT-qPCR cycling conditions included an initial uracil-DNA glycosylase incubation for 2 min at 25°C, reverse transcription for 15 min at 50°C, activation of the Taq enzyme for 2 min at 95°C, and a two-step cycling process involving 3 sec at 95°C and 30 sec at 55°C, repeated for a total of 45 cycles. A positive test result was determined by the presence of an exponential fluorescent curve intersecting the threshold within 40 cycles (cycle threshold [Ct] <40).

The quantification of PMMoV was executed using the TaqPath 1-Step RT-qPCR Master Mix, CG (Thermo Fisher Scientific) on a QuantStudios 7 Pro instrument. Each reaction was composed of 20 μL, including 5 μL of 4X Master Mix from Thermo Fisher Scientific, 0.5 μL of 10 μmol/L probe, 1.8 μL each of 10 μmol/L forward and reverse primers, 8.9 μL of nuclease-free water, and 2 μL of nucleic acid extract. The reagents were meticulously transferred into 96-well plates using pipettes and subsequently mixed by vortexing for 10 sec. The thermocycling conditions utilized in this study were as follows: incubation of uracil-DNA glycosylase for 2 min at 25°C, reverse transcription carried out for 15 min at 50°C, activation of the Taq enzyme for 10 min at 95°C, and a two-step cycling process consisting of 30 sec at 95°C followed by 1 min at 60°C, repeated for a total of 40 cycles.

In each RT-qPCR run, one positive PMMoV control and negative controls, comprising Mastermix and DNase/RNase-free water, were incorporated. The RT-qPCR reactions were carried out in triplicate, and the criteria for classifying a sample as positive included the requirement that all replicates produced positive results, with each individual replicate falling within the linear range of the standard curve. The N1 standard curve demonstrated a high level of efficiency, with a value of 94.669% (R2 = 1). The quantification of SARS-CoV-2 RNA was determined by calculating the average of three replicates of viral copies. The outputs of RT-qPCR were converted into units of copies per liter. In this study, the detection limit for SARS-CoV-2 and PMMoV was established at 20 and 10 copies per liter, respectively.

### DNA Isolation, 16S rRNA Gene Amplification, Sequencing

Before inclusion in the kit, quarter-sections of 0.45 µm and 0.22 µm nitrocellulose filters were prepared by flame-sterilizing a blade and using ethanol for sterilization. Genomic DNA extraction was then performed using the FastDNA Spin Kit for Soil (BIO101, Vista, CA, USA), strictly following the guidelines of manufacturer. Subsequent DNA purification utilized the SELECT-A-SIZE DNA Clean & Concentrator Kits (Zymo Research, Irvine, CA). The quality of the extracted DNA was assessed by determining the 260/280 and 260/230 ratios on a NanoDrop spectrophotometer (Thermo Fisher Scientific, Waltham, MA).

After confirming successful DNA extraction, Polymerase Chain Reaction (PCR) was conducted on 1–10 μL of the extracted DNA. DNA libraries were prepared following the methodology outlined by Caporaso et al. (2012). PCR amplification of the V4 region employed Phusion DNA polymerase (Master Mix; Thermo Fisher Scientific, Waltham, MA) and universal primers 515f and barcoded 806r, designed to anneal to both bacterial and archaeal sequences. A 12-bp barcode index on the reverse primer facilitated multiplexing for sequencing analysis.

Subsequently, amplicon quality and size were assessed using an Agilent Bioanalyzer (Agilent Technologies Santa Clara, CA). Following the protocol of manufacturer, the DNA amplicons were pooled and quantified with a NEBNext Library Quant Kit for Illumina (New England Biolabs, Ipswich, MA). Sequencing was performed using a MiSeq V2 kit on an Illumina MiSeq platform (Illumina, San Diego, CA).

Digital sequence data from the MiSeq underwent processing through the QIIME2 (v1.9) pipeline on a Linux Server (Caporaso et al. 2010). DADA2 within QIIME2 was employed for denoising, and fast-join facilitated the joining of paired-end sequences. Subsequent demultiplexing excluded sequences with a Phred score below 20, and UCHIME identified and removed chimeric sequences. Genus-level identification of sequences utilized the Silva database, with operational taxonomic units (OTUs) determined and sample populations normalized by total sequence count to ascertain the relative abundance of each OTU.

### Data Analysis

Statistical analyses were conducted using R version 4.2.3. Initially, samples were processed by rarefying OTU tables to the lowest library size across all samples in each student residence hall. Subsequently, we computed common l2-diversity metrics (Observed, ACE, Shannon, Simpson, InvSimpson, Fisher, Coverage, and PD) and l2-diversity metric (Bray-Curtis) using the R phyloseq package. To assess differences in l2-diversity metrics between groups, linear regression was employed, with semesters included as covariates. For the evaluation of differences in l2- diversity metrics between groups, nonmetric-multidimensional scaling (NMDS) was utilized, and *p* values for the comparison between groups were determined using permutational multivariate ANOVA models, which included semesters as covariates. A Pearson Correlation analysis was undertaken to explore the correlations between various parameters, including the relative abundance of enteric pathogen and potential pathogens, the relative abundance of enteric pathogen and potential pathogen in positive and negative SARS-CoV-2 samples, as well as the relative abundance of potential pathogen in positive and negative SARS-CoV-2 sewage samples.

## Results

### Concentration of SARS-CoV-2 in Raw Sewage

The 174 raw sewage samples included in this study were collected from 6 different dormitories in the same sewage network across the University of Tennessee, Knoxville (Figure 1). Figure 2 depicts the concentrations of SARS-CoV-2 from September 2020 to October 2021 within various high-density student residence halls. Over the sampling period, SARS-CoV-2 concentrations were consistently measured at different levels in the respective halls: 3.09±3.46 log10 copies/L in D1, 2.02±2.19 log10 copies/L in D2, 2.80±3.26 log10 copies/L in D3, 2.97±3.61 log10 copies/L in D4, 2.94±3.30 log10 copies/L in D5, and 2.36±2.71 log10 copies/L in D6.

**Figure 2.**
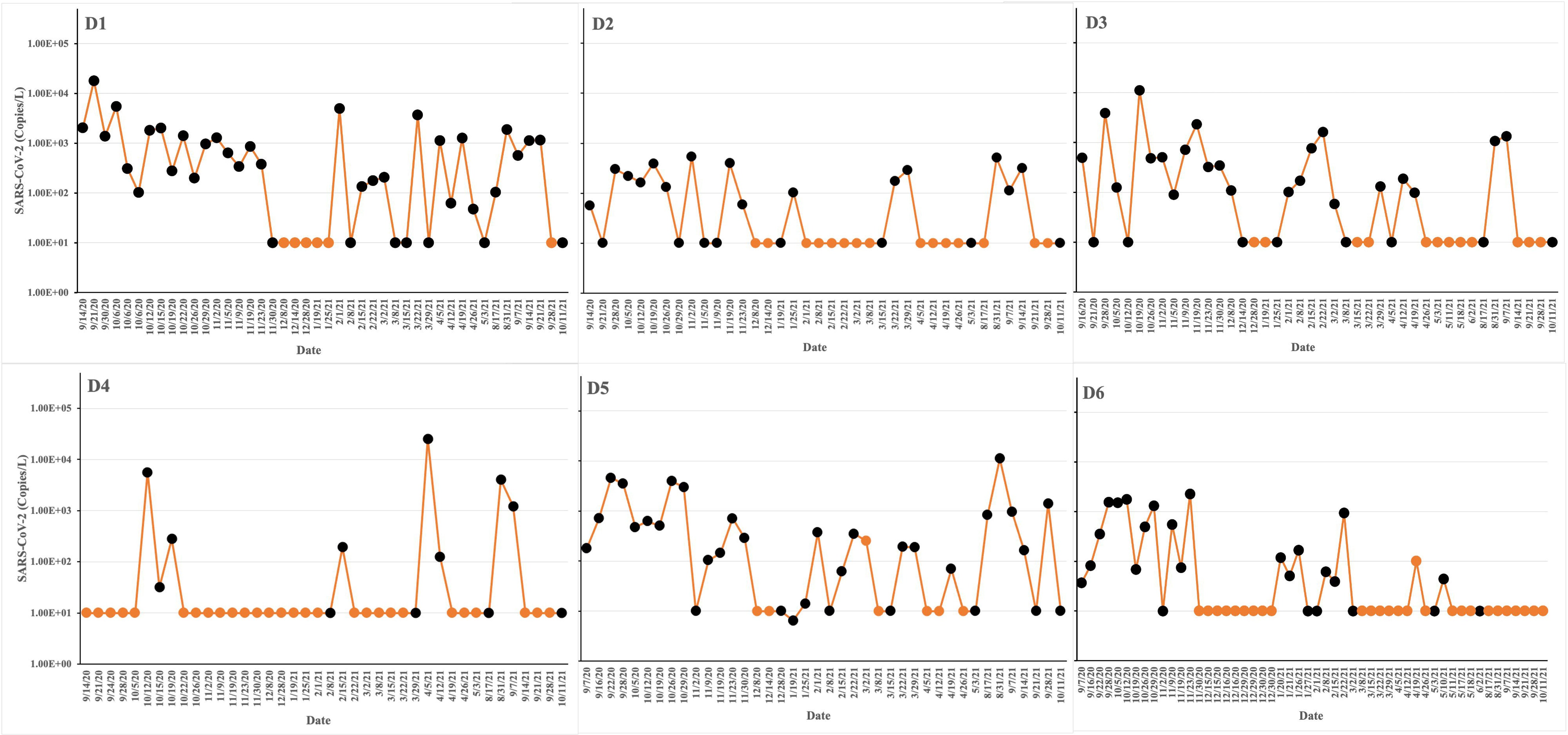
Experimental design and sampling points/times for microbiome sequencing. SARS- CoV-2 concentrations are indicated as yellow lines. The yellow points at 10 copies/L represent negative samples. The sequencing runs are indicated as black points. The virus load was estimated by qPCR in untreated wastewater from different dormitories: D1, D2, D3, D4, D5, and D6. The study was conducted from Sep 2020 to Oct 2021.

Furthermore, the positive rates, calculated by dividing the number of positive samples by the total number of samples and multiplying by 100%, varied across the halls. Specifically, the positive rates were 70% in D1, 39% in D2, 52% in D3, 20% in D4, 68% in D5, and 37% in D6 (Table 2). These results provide valuable insights into the dynamics of SARS-CoV-2 concentrations and positivity rates within high-density student residence halls during the specified timeframe.

**Table 2.** Raw wastewater data information for D1, D2, D3, D4, D5, and D6.

| Dorms | pH | Positive Rate | Relative abundance of potential Pathogen |  | 602 |
| --- | --- | --- | --- | --- | --- |
|  |  |  | Positive SARS-CoV-2 Sample | Negative SARS-CoV-2 Sample |  |
| D1 | 6.71-9.08 | 70.00% | 40.04% | 40.35% |  |
| D2 | 6.83-8.27 | 39.00% | 21.80% | 29.37% |  |
| D3 | 6.51-8.97 | 52.00% | 17.58% | 13.88% |  |
| D4 | 6.27-9.01 | 20.00% | 30.63% | 24.11% |  |
| D5 | 6.38-8.95 | 68.00% | 43.49% | 45.17% |  |
| D6 | 5.72-8.63 | 37.00% | 41.30% | 45.64% |  |

### Characteristics of the predominant flora in different dormitories

#### Characterization on phylum, family, and genus level

The sequences extracted from the samples underwent comprehensive analysis, resulting in the classification of data into 56 phyla, 145 classes, 315 orders, 548 families, and 1170 genera.

Figure 3 illustrates the relative abundances of the top 10 phyla across six dormitories, revealing notable variations. The phylum Bacteroidetes was identified as the most abundant across all sampling sites, with relative abundance ranging from 46.1% to 26.9%. Firmicutes emerged as the second most abundant phylum in dormitories 1, 2, 3, and 4, with relative abundance varying from 35.2% to 14.8%. Meanwhile, Proteobacteria emerged as the second most abundant phylum in dormitories 5 and 6, with relative abundance ranging from 41.3% to 25.4%. Additionally, it was observed that dormitories 2 and 3 have the same four highest abundance phyla of Bacteroidetes, Firmicutes, Proteobacteria, and Spirochaetota. Similarly, Dormitories 5 and 6 have the same four highest abundance phyla of Bacteroidetes, Proteobacteria, Firmicutes, and Campilobacterota.

**Figure 3.**
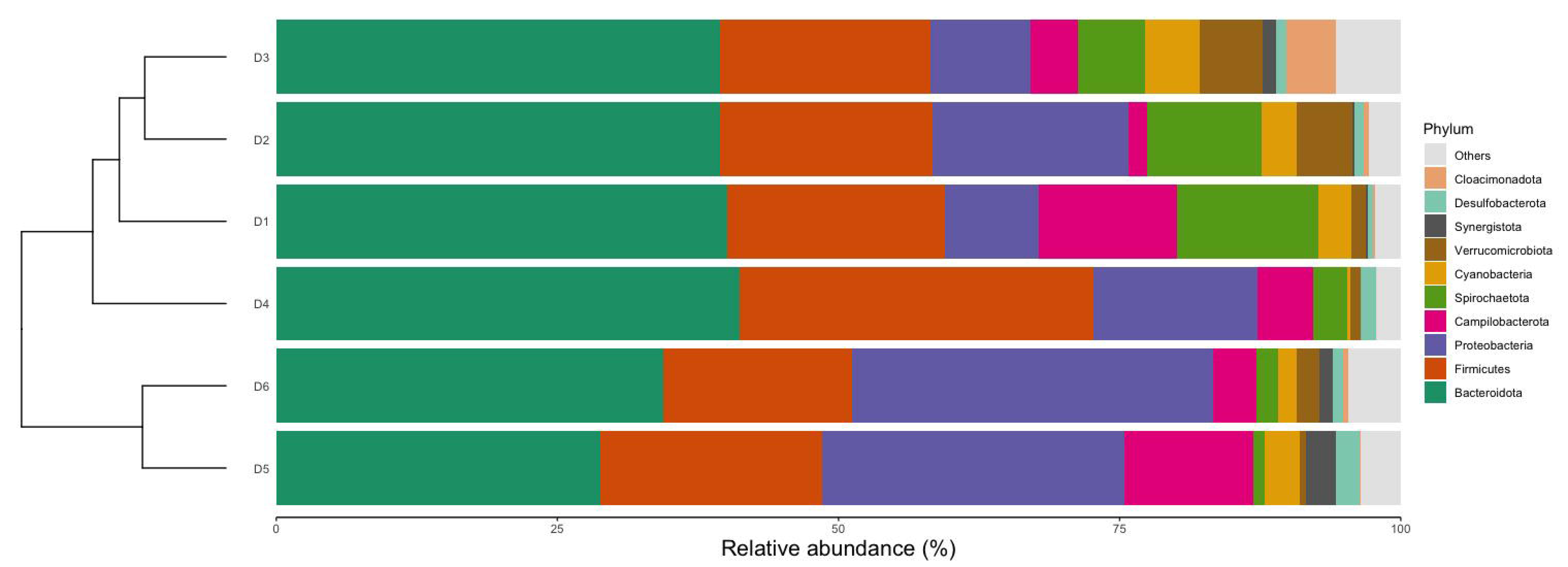
Relative abundances of the top 10 dominant phyla in 6 dormitories.

Figure 4 highlights the 10 most dominant families, showcasing significant differences among the six dormitories. Paludibacteraceae and Spirochaetaceae emerged as the most dominant families, with varying relative abundance in D1, D2, and D3. Interestingly, Bacteroidaceae took precedence in D4, D5, and D6, with distinct relative abundance up to 12.6%.

**Figure 4.**
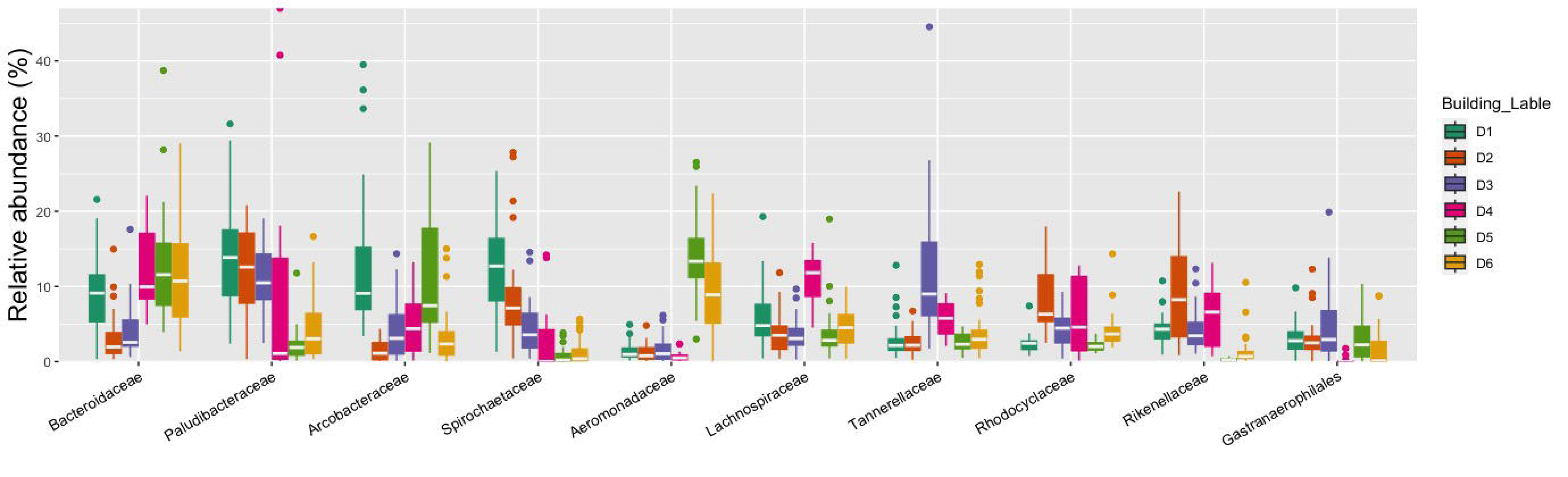
Relative abundances at family levels for six dormitories.

The relative abundances of the top 50 genera detected in all dormitory samples are shown in Figure 5. Moreover, many typical gut bacteria were also found at very high levels in the sewage such as Bacteroides, Acinetobacter, Prevotella, Pseudomonas, Blautia, Faecalibacterium, Ruminococcus, Dorea, and others, corresponding to ranks 1, 8, 10, 11, and 21 within the top 50 genera in Figure 5 (Furet et al. 2009, Cai et al. 2014, Bäckhed et al. 2015, Do et al. 2019).

**Figure 5.**
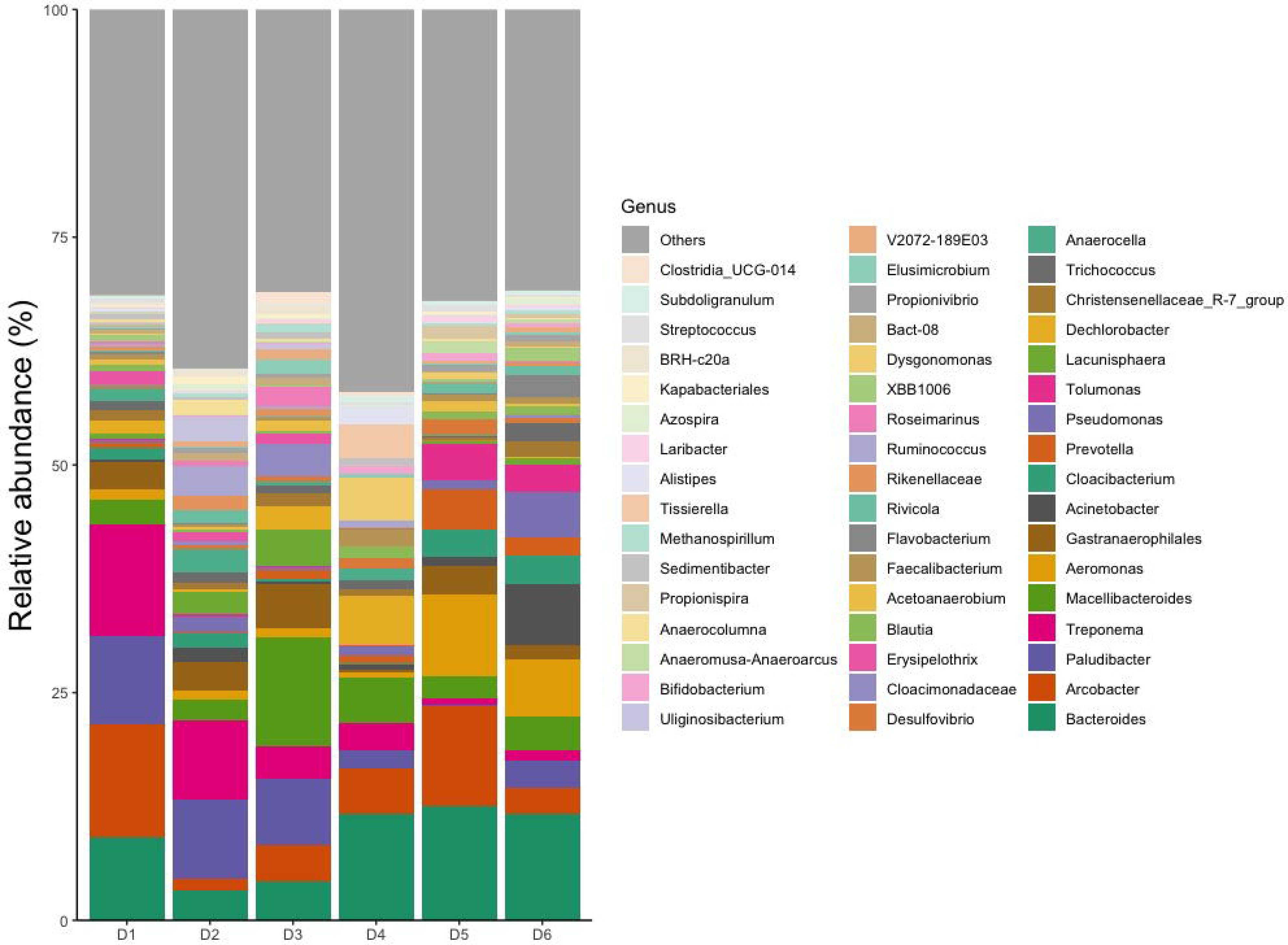
Relative abundances of top 50 genera.

Among the top 50 genera, 14 genera (33.52%) were identified as potential pathogens, including Bacteroides, Arcobacter, Treponema, Aeromonas, Acinetobacter, Prevotella, Pseudomonas, Erysipelothrix, Faecalibacterium, Flavobacterium, Ruminococcus, Bifidobacterium, Laribacter, and Streptococcus (Cai and Zhang 2013, Cai et al. 2014, Do et al. 2019, Oluseyi Osunmakinde et al. 2019, Poopedi et al. 2023).

In the examination of the top 50 genera, dormitories D1 to D6 exhibited varying relative abundance of potential pathogens, with 13 (40%), 12 (23%), 11 (18%), 12 (28%), 14 (45%), and 14 (41%) genera recognized as such, respectively (Figure 5). Notably, a substantial number of these potential pathogens displayed an increased relative abundance in samples from D1, D5, and D6 compared to other sites. Among the detected enteric pathogens in the top 50 genera were *Arcobacter*, *Aeromonas*, and *Laribacter*, with total relative abundances of 13.61%, 2.37%, 5.45%, 5.64%, 20.82%, and 9.01% from D1 to D6, respectively. *Arcobacter* and *Aeromonas* were identified across all six dormitories, while *Laribacter* was exclusively found in D3, D5, and D6. Furthermore, an observation revealed a correlation between the relative abundance of enteric pathogen and potential pathogens (Pearson Correlation = 0.842, *p* = 0.018). Additionally, *Mycobacterium*, the most prevalent respiratory tract-associated pathogen, contributed from 0.02% to 0.15% of the total bacterial community across all six dormitories, respectively.

### Diversity of bacterial communities

The analysis of the microbiota communities within the collected wastewater samples revealed significant distinctions across all sampled locations (Figure 6). At the species level, dormitory 6 (D6) exhibited the highest count of exclusive taxa, totaling 1206, while the other dormitories (D1 to D5) displayed varying counts of exclusive taxa, ranging from 546 to 1081 species. It is noteworthy that a core microbiome consisting of 286 bacterial species was consistently observed across all sampled dormitories.

**Figure 6.**
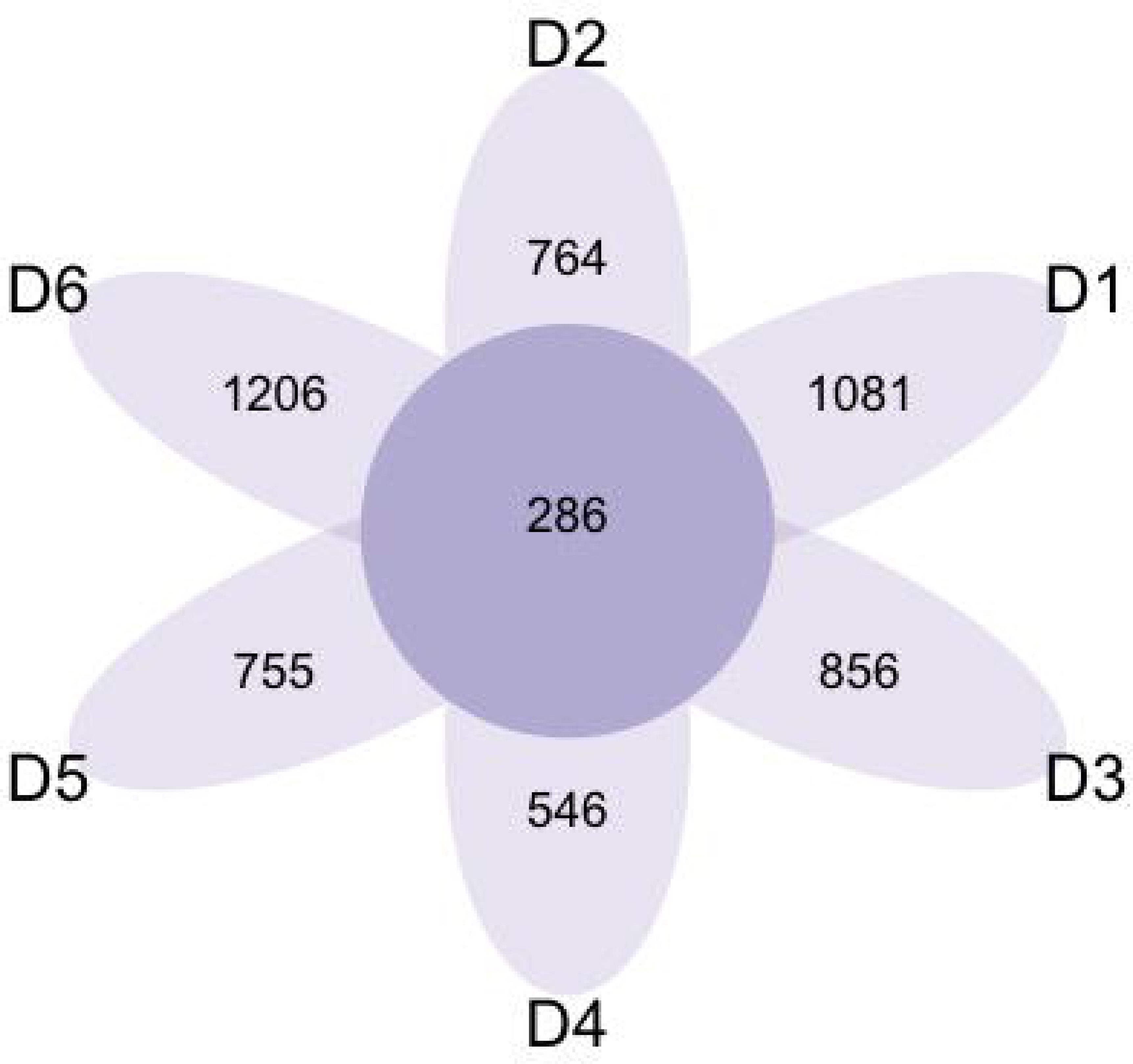
Venn diagram of exclusives and shared bacteria among the 6 dormitories.

Considerable distinctions were identified in microbiota communities within the collected wastewater from all sampled locations, as illustrated in Figure 6. Notably, at the species level, D6 demonstrated the highest count of exclusive taxa, totaling 1206. Conversely, the other dormitories (D1 to D5) exhibited diverse counts of exclusive taxa, with 1081, 764, 856, 546, and 755 species, respectively. Notably, a core microbiome was observed with 286 bacterial species.

Alpha diversity analysis was employed to assess the diversity and richness of bacterial communities within the microbiome of six dormitories (Figure 7). A comprehensive comparison among the six dormitories was conducted using linear regression models, with semester serving as a covariate. The results showed statistically significant differences in bacterial diversity across the dormitories.

**Figure 7.**
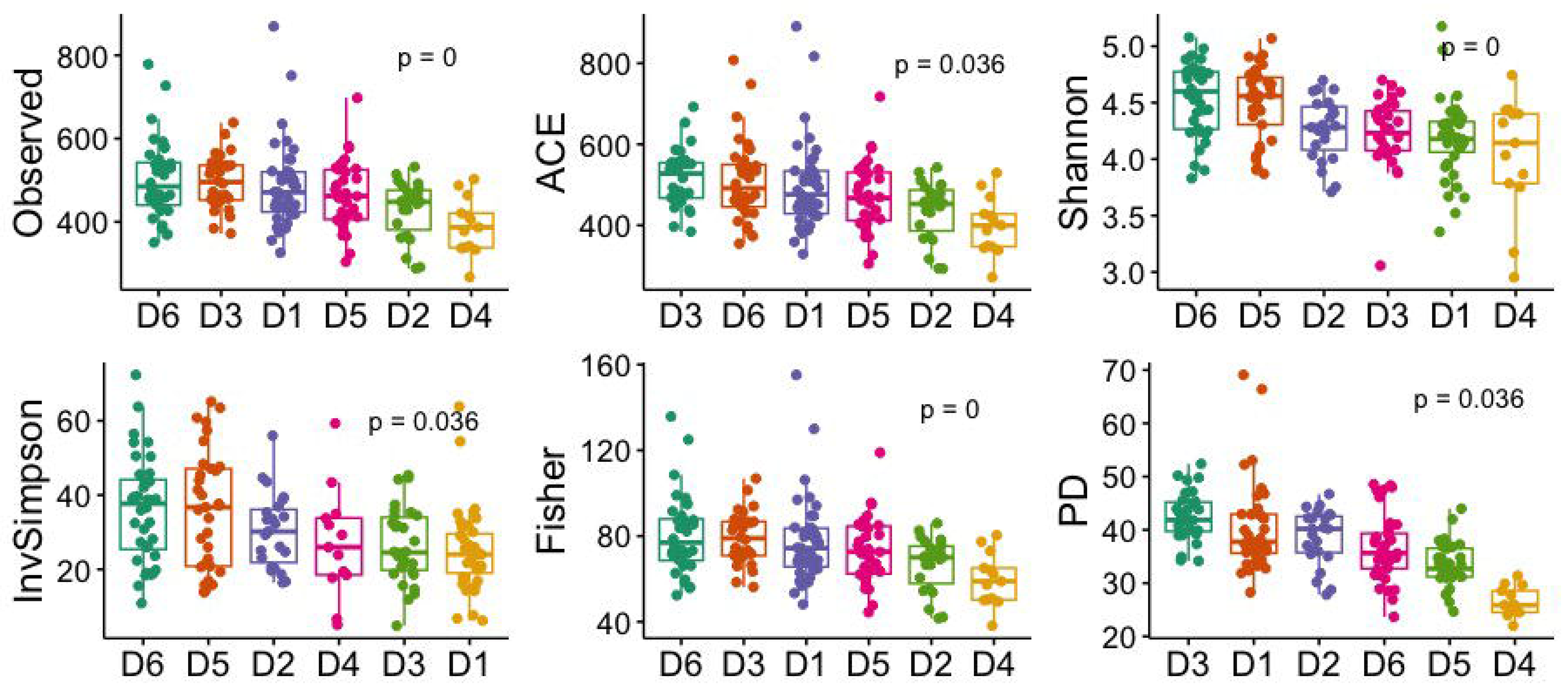
Diversity index in 6 dormitories. The box-and-whisker plots show the mean (diamond), median (middle bar), first quartile (lower bar), third quartile (upper bar), minimum observation above the lowest fence (lower whisker), and maximum observation below the upper fence (upper whisker) of common l2-diversity metrics for each group. The P values for the comparison between groups using linear regression models including semester as covariate is also shown.

Figure 8 depicts the clustering of beta diversity, assessed through Bray-Curtis distance metrics, among the six dormitories. A permutational multivariate ANOVA model, which included semester as a covariate, demonstrated significant differences in the measured β-diversity metrics between groups (*p* < 0.05).

**Figure 8.**
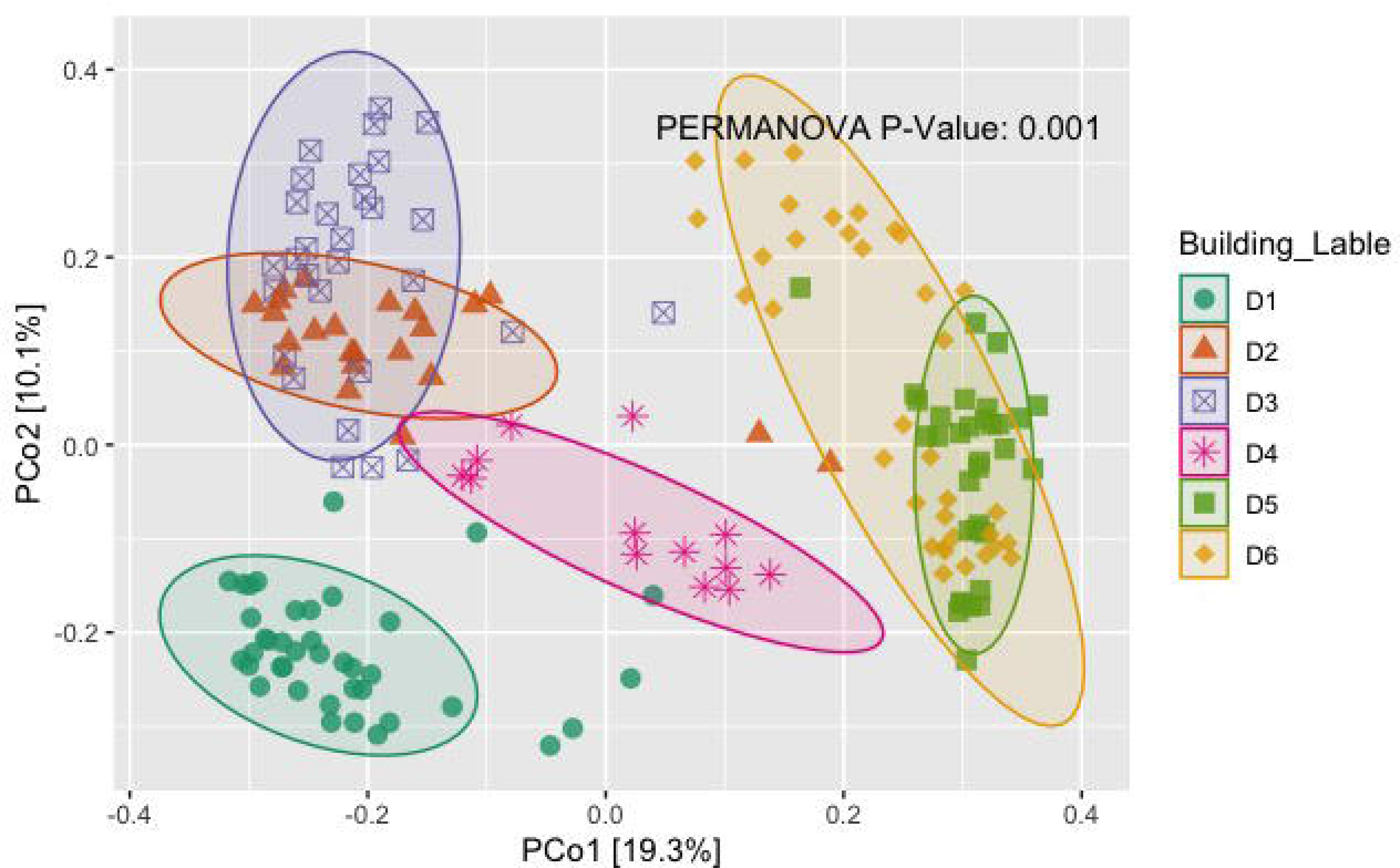
The scatter plots show each participant’s microbial community composition (small circles) by group, as well as each group’s centroid (large circles) and 95% CI ellipses. The scatter plots were generated using Principal Coordinates Analysis (PCoA) ordination based on common β -diversity metrics. For ease of visualization, only 2 dimensions were used. The P values for the comparison between groups using permutational multivariate ANOVA models including semester as covariate is also show.

### Characteristics of the predominant flora in positive and negative samples Characterization on phylum, family, and genus level

The microbial composition of various sampling sites was analyzed to determine the abundance of specific phyla and family, with a focus on the impact of COVID-19 status on the results.

These results showed that the microbial composition of different dormitory locations remains consistent at the top 10 dominant phyla and family, regardless of the COVID-19 status (Figure 9, 10).

**Figure 9.**
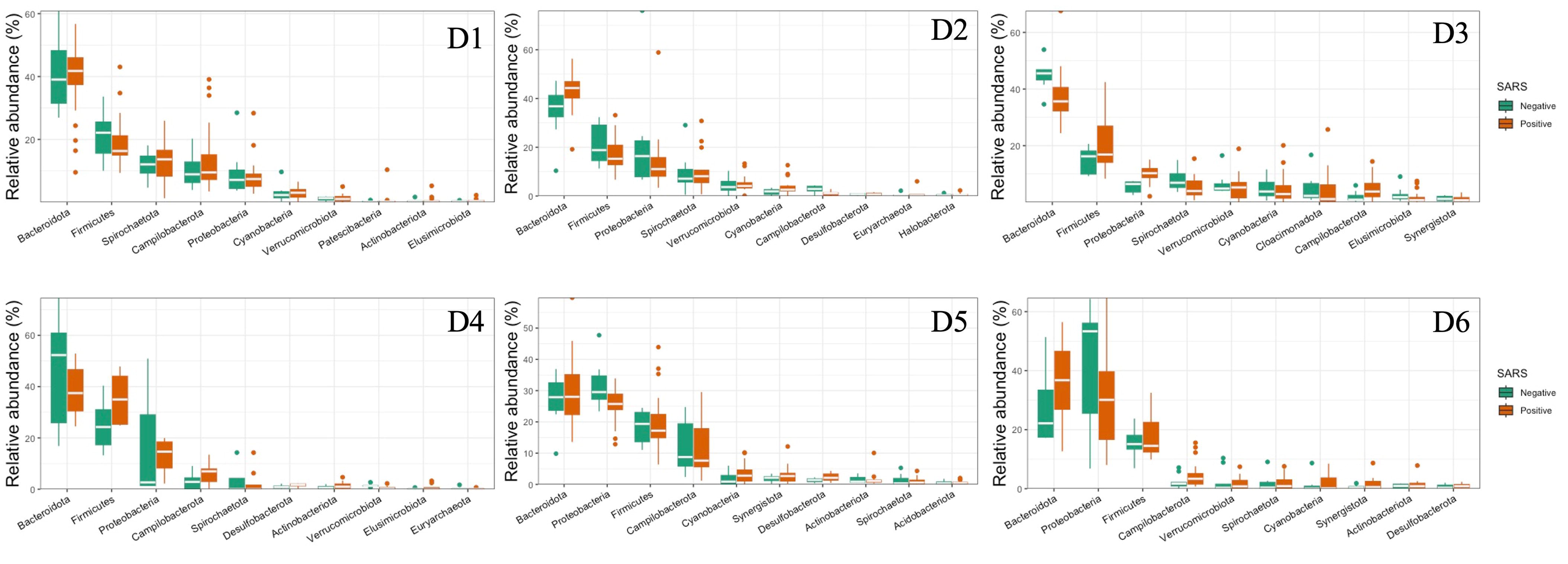
Relative abundances of the top 10 dominant phyla in 6 dormitories with positive and negative SARS-CoV-2 samples.

**Figure 10.**
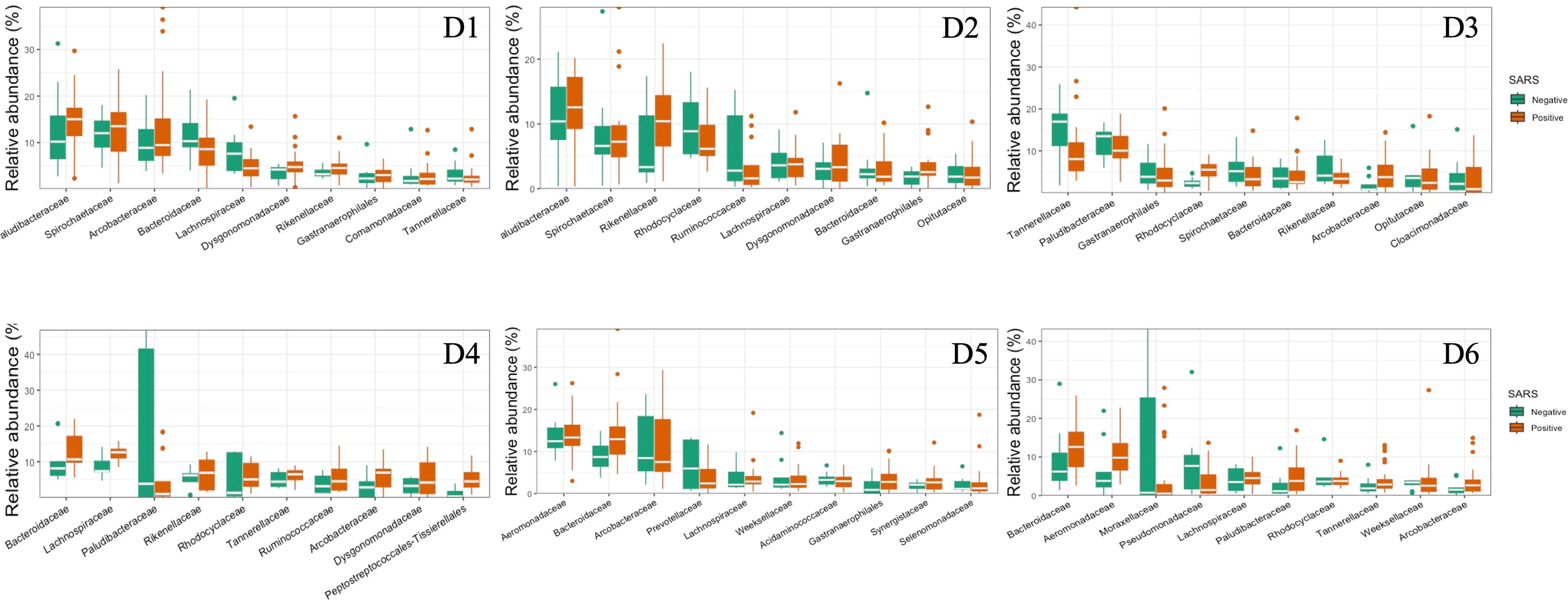
Relative abundances of the top 10 dominant family in 6 dormitories with positive and negative SARS-CoV-2 samples.

The current study, encompassing multiple dormitories, unveiled several noteworthy distinctions in the relative abundances of microbial families (Figure 11). Dormitory 1 showed a distinction between the families Lachnospiraceae and Streptococcaceae. Similarly, dormitory 3 exhibited significant differences in the relative abundances of Arcobacteraceae, Peptostreptococcaceae, Rhodocyclaceae, and V2072-189E03 between COVID-19 positive and negative samples. In dormitory 4, Peptostreptococcales-Tissierellales displayed significant variation in relative abundances between the two sample groups. In dormitory 5, Desulfovibrionaceae demonstrated a significant difference in relative abundances based on COVID-19 status. Lastly, in dormitory 6, significant differences were observed in the relative abundances of Aeromonadaceae and Paludibacteraceae between COVID-19 positive and negative samples. These findings underscore the potential impact of COVID-19 on specific microbial families within the microbiota of different dormitories, providing valuable insights into the nuanced variations in microbial composition associated with the viral infection.

**Figure 11.**
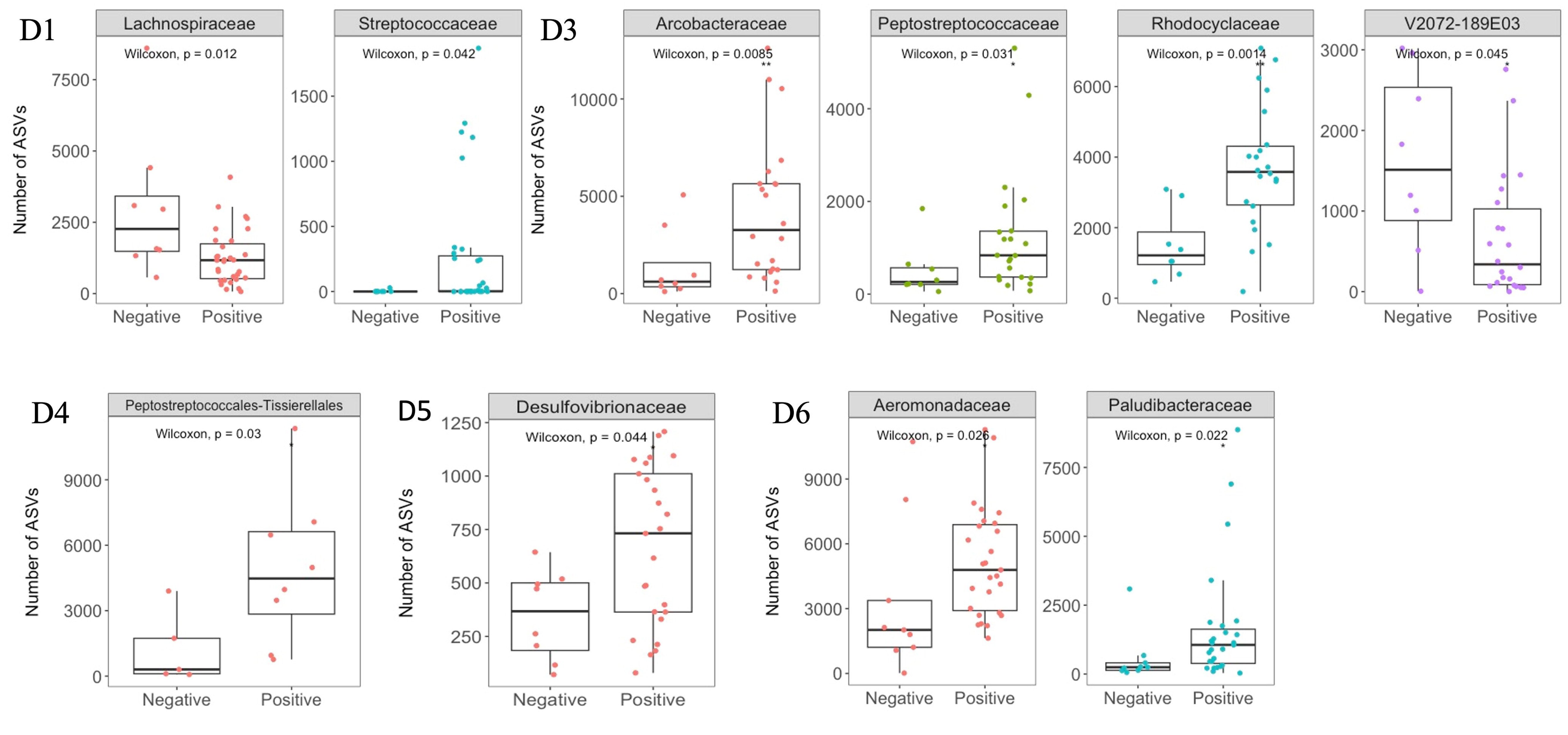
Significant changes at family levels with positive and negative SARS-CoV-2 samples in 6 dormitories.

The LEfSe analysis was employed to discern and differentiate the microbiome composition between samples that tested positive and negative for COVID-19 across the six dormitories (Figure S1). Interestingly, no universal biomarkers were identified across all six dormitories. Instead, distinct biomarkers were exclusively found in dormitories 3, 4, and 5, suggesting unique microbial signatures associated with COVID-19 status in these specific dormitory environments.

The present study aimed to analyze the relative abundance of potential pathogen in both SARS-CoV-2 positive and negative samples of the top 50 genera (Figure 12). As depicted in Table 2, the results revealed a noteworthy correlation between the relative abundance of potential pathogen in positive and negative samples, with a Pearson Correlation coefficient of 0.918 (*p* = 0.010). Additionally, the study found a significant correlation between the relative abundance of enteric pathogen and potential pathogen in positive SARS-CoV-2 samples (Pearson Correlation = 0.817, *p* = 0.024), irrespective of relative abundance of potential pathogen in negative SARS-CoV-2 samples.

**Figure 12.**
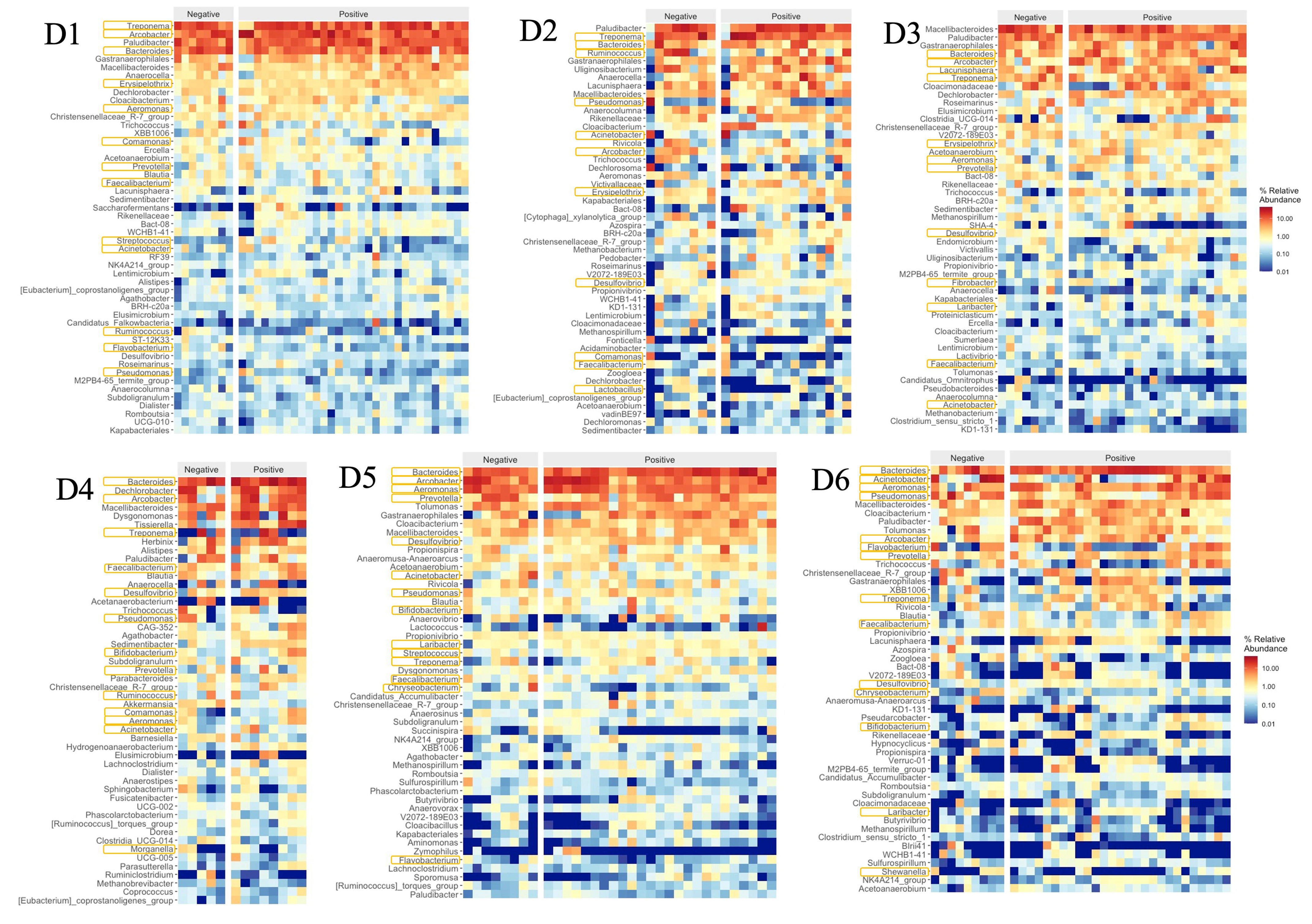
Relative abundances of top 50 genera and potential pathogens with positive and negative SARS-CoV-2 samples in 6 dormitories. The genera are listed from the highest relative abundance (top) to the least relative abundance (bottom). The pathogens are marked with an orange box around their name.

### Diversity of bacterial communities

Our study conducted a comprehensive analysis of samples from different locations, with a specific focus on viral quantification to distinguish the differences between exclusive and shared species in wastewaters tested for SARS-CoV-2. We found that samples testing positive for SARS-CoV-2 demonstrated a higher diversity of taxa compared to their negative counterparts (Figure 13). The analysis highlighted that exclusive species were most prominently represented in positive samples for SARS-CoV-2 collected from D1 at 31.87%, while D3 exhibited the lowest representation at 18.99%. Conversely, negative samples for SARS-CoV-2 were associated with exclusive bacterial species in wastewater collected from D3 (18.24%), with D1 displaying the lowest representation at 8.49%. Despite the SARS-CoV-2 status, the analysis further indicated a low representativity for exclusive bacteria found in other dormitories.

**Figure 13.**
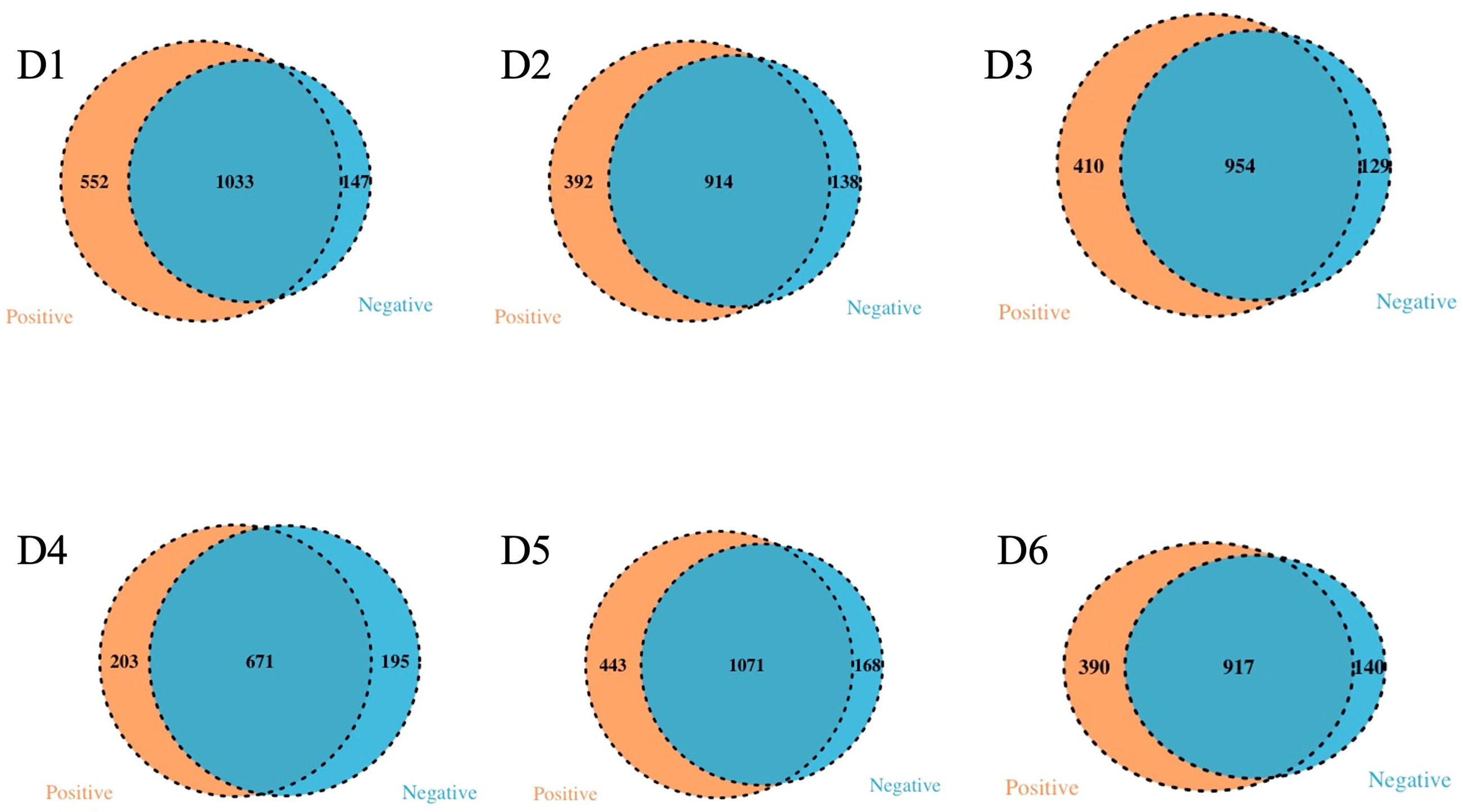
Venn diagram of exclusives and shared bacteria with positive and negative SARS- CoV-2 samples in the 6 dormitories.

An observation revealed a correlation between the positive rate of sampling sites and the relative abundance of exclusive species in positive samples (Pearson Correlation = 0.771, *p* = 0.036).

Additionally, the presence of 1033, 914, 954, 671, 1071, and 917 taxa in both SARS-CoV-2 positive and negative samples from D1 to D6, respectively. These findings collectively contribute to our understanding of the microbial dynamics associated with SARS-CoV-2 in wastewater samples across different dormitory locations.

The l2-diversity of the microbiome across all locations exhibited a general trend of being higher in negative samples compared to positive samples. Specifically, the observed species index showed a significant difference in D5 and D6 (p < 0.05, Figure 14), as determined through linear regression models that incorporated semester as a covariate. Notably, significant differences in the measured β-diversity metrics were discerned in D3 and D6 between groups (p < 0.05 for the Bray-Curtis indices, using permutational multivariate ANOVA with semester as a covariate), as illustrated in Figure 15.

**Figure 14.**
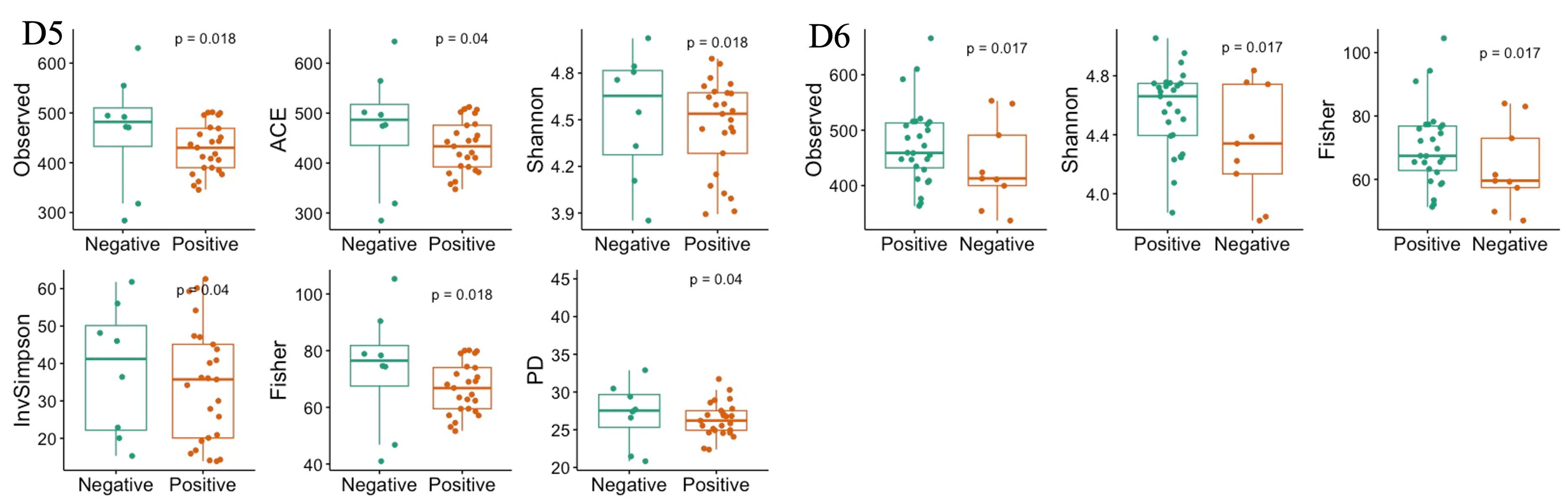
Diversity index with significant difference between the positive and negative SARS- CoV-2 samples in 6 dormitories. The box-and-whisker plots show the mean (diamond), median (middle bar), first quartile (lower bar), third quartile (upper bar), minimum observation above the lowest fence (lower whisker), and maximum observation below the upper fence (upper whisker) of common l2-diversity metrics just for significant group. The P values for the comparison between groups using linear regression models including semester as covariate is also shown.

**Figure 15.**
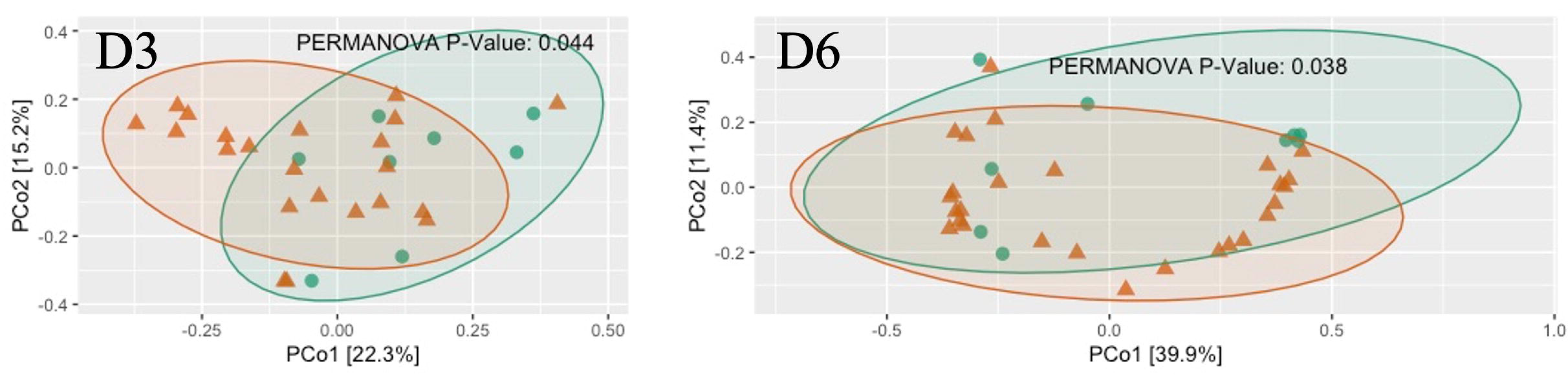
The scatter plots show each participant’s microbial community composition (small circles) by D4 and D6, as well as their centroid (large circles) and 95% CI ellipses. The scatter plots were generated using Principal Coordinates Analysis (PCoA) ordination based on common b-diversity metrics. For ease of visualization, only 2 dimensions were used. The P values for the comparison between groups using permutational multivariate ANOVA models including semester as covariate is also shown.

## Discussion

The identified variations at the phylum, family, and genus levels across the six dormitories shed light on the geographic differences in bacterial composition in this study (Figure 3, 4, and 5).

The analysis revealed two clusters of community types, as illustrated in Figure 3. Dormitories 2, 3, 1, and 4, organized by the closer relationship of bacterial phyla in each building, exhibited similar dominance patterns in these phyla, while D5 and D6 exhibited comparable compositions. The spatial arrangement depicted in the map (Figure 1) highlights that D1, D2, and D4 are in proximity, D5 and D6 are likewise nearby, and D3 is closer to D5 and D6. This spatial variation suggests a potential impact of geographic factors on the microbial composition in different dormitories. This observation aligns with the study by Fierer et al. (2022) finding five clusters of 17 different locations, revealing no strong relationship with the distance between sampling locations.

The significant alpha and beta diversity further underscore pronounced geographical variations in microbial communities in this study, aligning with Fierer et al. (2022) findings. Their emphasis on independently considering spatial variations when assessing the wastewater microbiome highlights the need to account for the influence of location on microbial diversity. Their research identified geographic variations in bacterial composition unrelated to sewer material, sewer depth, or resident human population on the campus. They attributed these variations to sample pH, with total suspended solids concentrations and sample volume playing a lesser role. This pH correlation aligns with studies by Fujii et al. (2012) and Lindström et al. (2005), which demonstrated the close association between pH and shifts in bacterial community composition in aquatic environments. Despite the detected variations in bacterial composition across dormitories in our study, the pH did not exhibit significant changes. Future research could explore specific factors such as organic carbon or nutrient concentrations to better understand the observed geographic variations in microbial communities.

The analysis of the microbial community in raw sewage yielded results consistent with previous research, indicating the influence of the human gut bacterial community on the bacterial profile in raw sewage. Specifically, the phyla Bacteroidota was identified as the most abundant and variable across samples, aligning with findings from Arumugam et al. (2011). However, a study by Cai et al. (2014) reported Firmicutes as the most dominant phylum in influent samples, asserting its alignment with the human microbiome composition. The findings of may clarify this discrepancy Turnbaugh et al. (2006) and Clemente et al. (2012), indicating that the gut microbiota typically showcases dominance of bacteria, particularly from the Bacteroidota and Firmicutes divisions. Furthermore, Huttenhower et al. (2012) revealed that gut microbiota relationships were characterized by inverse associations with Bacteroidota, varying from dominance in some subjects to a minority in others with a greater diversity of Firmicutes. These nuanced observations highlight the intricate dynamics of the human gut microbiota and underscore the pivotal roles played by Bacteroidota and Firmicutes in shaping microbial profiles observed in raw sewage.

The primary objective of our study was to investigate potential changes in wastewater microbiomes during the pandemic, considering the influence of the human gut bacterial community on the bacterial profile in raw sewage. This investigation was prompted by existing literature highlighting significant alterations in fecal microbiomes among individuals with COVID-19 (Gu et al. 2020, Zuo et al. 2020, Yeoh et al. 2021). The significant differences in bacterial composition observed across the six dormitories prompted a recommendation for separate analyses of the 16S rRNA data for each dormitory. This approach aims to mitigate biases that may arise when combining data from diverse dormitory settings. The prominent representation of exclusive species in positive samples for SARS-CoV-2 were found across all six dormitories supports the findings of Gallardo-Escárate et al. (2021). Moreover, the observed trend of higher l2-diversity in the microbiome of negative samples compared to positive samples across some locations echoes the results reported by Gu et al. (2020) and Yeoh et al. (2021), who documented a significant decrease in gut microbiota diversity and abundance in COVID-19 patients relative to healthy individuals.

The observed correlation between the relative abundance of enteric pathogen and potential pathogens at sampling sites adds a significant layer of understanding in the context of COVID- 19, particularly highlighting the notable association between the relative abundance of enteric pathogen and potential pathogen in positive SARS-CoV-2 samples. The presence of three enteric genera, namely, *Arcobacter*, *Aeromonas*, and *Laribacter*, in our study, commonly residing in the human intestines and potentially utilizing pathogenic mechanisms to induce gastrointestinal tract infections, emphasizes the relevance of these microbes in the sewage context during the pandemic. Notably, the *Aeromonas* genus ranked as the third leading cause of diarrhea after Campylobacter and Salmonella ￼, exhibited a notably high abundance exclusively in D5 (9.09%) and D6 (6.14%) compared to other dormitories, where the abundance ranged from 0.66% to 1.14%. Additionally, two *Arcobacter* species, *A. butzleri, and A. cryaerophilus,* are considered emerging pathogens posing threats to human health*, adding* depth to discussing potential pathogenic risks in the sewage microbiome. Additionally, the genus *Laribacter*, represented by the species *L. hongkongensis*, known for its associations with traveler gastroenteritis and diarrhea (Beilfuss et al. 2015), further contributes to understanding the microbial landscape in the context of COVID-19.

In the context of the ongoing discourse surrounding COVID-19, an emerging respiratory infectious disease, the investigation into the presence of *Mycobacterium*, a medically significant respiratory tract-associated pathogen, within sewage systems has garnered attention. Notably, this scrutiny extends across six dormitories, revealing a discernibly lower total abundance of *Mycobacterium* in sewage than the prevalent genera identified in the samples. The quantification of *Mycobacterium* in our samples aligns with findings from previous studies, providing a basis for comparative analysis. Cai and Zhang (2013) reported an overall abundance of *Mycobacterium* in influent and effluent samples that remained below the threshold of 0.02%.

Numberger et al. (2019) the genus Mycobacterium was observed exclusively in October effluent samples with a relative abundance of less than 0.02%. The 16S rRNA gene sequences analysis in our work determined the presence of the bacterial genera but not species. These genera may contain both pathogenic and non-pathogenic species. Therefore, the identification of pathogens requires further study.

Our study did not unveil a significant universal biomarker distinguishing positive from negative SARS-CoV-2 sewage samples across all sampling locations. This contrasts with the findings of Gu et al. (2020), who identified five biomarkers to differentiate between COVID-19 patients and healthy individuals. It is essential to note that the absence of SARS-CoV-2 detection in certain patients may not necessarily signify a complete recovery of their gut microbiota. The restoration of microbial communities may require an extended period, even when SARS-CoV-2 is not detectable. This aligns with the observations of Zhang et al. (2023), who documented persistent dysbiosis for months after the clearance of the virus. Individuals recovered from COVID-19, when compared to healthy controls, exhibited reduced bacterial diversity and richness at 3 months. This reduction was accompanied by a lower abundance of beneficial commensals and a higher abundance of opportunistic pathogens. Hence, the significant correlation in the relative abundance of potential pathogen between positive and negative SARS-CoV-2 sewage samples in our study may be attributed to the lingering effects of microbial dysbiosis observed in COVID- 19 recovery.

## Conclusion

In conclusion, our study provides valuable insights into the raw sewage microbiota as a reflection of the gut microbiota during the COVID-19 pandemic and its potential association with fecal SARS-CoV-2 shedding. The observed significant differences in raw sewage microbial communities across all sampling sites and the prominent representation of exclusive species in positive samples for SARS-CoV-2 emphasize the potential of sewage microbiota as an indicator of viral shedding. Positive samples for SARS-CoV-2 exhibited a significant reduction in bacterial diversity, highlighting the impact of the virus infection on microbial composition.

These findings introduce a novel and targeted approach for modulating sewage microbiota, specifically linked to gastrointestinal manifestations, as a strategy for monitoring and predicting the presence of SARS-CoV-2 in raw sewage.

While our analysis did not uncover a significant universal biomarker distinguishing positive and negative SARS-CoV-2 raw sewage samples, the observed noteworthy correlation in the relative abundance of potential pathogens between these samples suggests a potential connection to the enduring effects of microbial dysbiosis during the recovery phase of COVID-19. Moreover, the identified correlation between the relative abundance of enteric pathogens and potential pathogens at sampling sites adds a significant dimension to our understanding of COVID-19, particularly in the context of the substantial correlation in positive SARS-CoV-2 samples. These findings underscore the importance of monitoring enteric pathogens in raw wastewater surveillance systems to comprehend the potential spread of COVID-19 and other infectious diseases. Such insights carry crucial implications for public health monitoring and management strategies.

## Data Availability

All data produced in the present study are available upon reasonable request to the authors

## Acknowledgment

The authors would like to thank the Office of Research & Engagement of the University of Tennessee, Knoxville, for providing funding for the project.

---

Figure S1. Histograms of linear discriminant analysis (LDA) effect size (LEfSe) comparison between positive and negative SARS-CoV-2 samples microbiota at the genus level in D3, D4 and D5. Log-level changes in LDA score are displayed on the x axis

## References

1. Aktaş, B. and Aslim, B. (2020). “Gut-lung axis and dysbiosis in COVID-19.” Turkish journal of biology 44(7): 265–272 DOI: 10.3906/biy-2005-102.

2. Arumugam, M., Raes, J., Pelletier, E., Le Paslier, D., Yamada, T., Mende, D. R., Fernandes, G. R., Tap, J., Bruls, T., Batto, J.-M., Bertalan, M., Borruel, N., Casellas, F., Fernandez, L., Gautier, L., Hansen, T., Hattori, M., Hayashi, T., Kleerebezem, M., Kurokawa, K., Leclerc, M., Levenez, F., Manichanh, C., Nielsen, H. B., Nielsen, T., Pons, N., Poulain, J., Qin, J., Sicheritz-Ponten, T., Tims, S., Torrents, D., Ugarte, E., Zoetendal, E. G., Wang, J., Guarner, F., Pedersen, O., de Vos, W. M., Brunak, S., Doré, J., Antolín, M., Artiguenave, F., Blottiere, H. M., Almeida, M., Brechot, C., Cara, C., Chervaux, C., Cultrone, A., Delorme, C., Denariaz, G., Dervyn, R., Foerstner, K. U., Friss, C., van de Guchte, M., Guedon, E., Haimet, F., Huber, W., van Hylckama-Vlieg, J., Jamet, A., Juste, C., Kaci, G., Knol, J., Kristiansen, K., Lakhdari, O., Layec, S., Le Roux, K., Maguin, E., Mérieux, A., Melo Minardi, R., M’Rini, C., Muller, J., Oozeer, R., Parkhill, J., Renault, P., Rescigno, M., Sanchez, N., Sunagawa, S., Torrejon, A., Turner, K., Vandemeulebrouck, G., Varela, E., Winogradsky, Y., Zeller, G., Weissenbach, J., Ehrlich, S. D., Bork, P. and Meta, H. I. T. C. (2011). “Enterotypes of the human gut microbiome.” Nature 473(7346): 174–180 DOI: https://10.1038/nature09944.

3. Ash, K. T., Li, Y., Alamilla, I., Joyner, D. C., Williams, D. E., McKay, P. J., Green, B. M., Iler, C., DeBlander, S. E., North, C. M., Kara-Murdoch, F., Swift, C. M. and Hazen, T. C. (2023). “SARS-CoV-2 raw wastewater surveillance from student residences on an urban university campus.” Frontiers in Microbiology 14 DOI: 10.3389/fmicb.2023.1101205.

4. Bäckhed, F., Roswall, J., Peng, Y., Feng, Q., Jia, H., Kovatcheva-Datchary, P., Li, Y., Xia, Y., Xie, H. and Zhong, H. (2015). “Dynamics and stabilization of the human gut microbiome during the first year of life.” Cell host & microbe 17(5): 690–703 DOI: 10.1016/j.chom.2015.04.004.

5. Beilfuss, H. A., Quig, D., Block, M. A. and Schreckenberger, P. C. (2015). “Definitive identification of Laribacter hongkongensis acquired in the United States.” Journal of Clinical Microbiology 53(7): 2385–2388 DOI: 10.1128/jcm.00539-15.

6. Cai, L., Ju, F. and Zhang, T. (2014). “Tracking human sewage microbiome in a municipal wastewater treatment plant.” Applied microbiology and biotechnology 98: 3317–3326 DOI: 10.1007/s00253-013-5402-z.

7. Cai, L. and Zhang, T. (2013). “Detecting Human Bacterial Pathogens in Wastewater Treatment Plants by a High-Throughput Shotgun Sequencing Technique.” Environmental Science & Technology 47(10): 5433–5441 DOI: 10.1021/es400275r.

8. Caporaso, J. G., Kuczynski, J., Stombaugh, J., Bittinger, K., Bushman, F. D., Costello, E. K., Fierer, N., Peña, A. G., Goodrich, J. K. and Gordon, J. I. (2010). “QIIME allows analysis of high-throughput community sequencing data.” Nature methods 7(5): 335–336 DOI: https://doi-org.utk.idm.oclc.org/10.1038/nmeth.f.303.

9. Caporaso, J. G., Lauber, C. L., Walters, W. A., Berg-Lyons, D., Huntley, J., Fierer, N., Owens, S. M., Betley, J., Fraser, L. and Bauer, M. (2012). “Ultra-high-throughput microbial community analysis on the Illumina HiSeq and MiSeq platforms.” The ISME journal 6(8): 1621–1624 DOI: https://doi-org.utk.idm.oclc.org/10.1038/ismej.2012.8.

10. Clemente, J. C., Ursell, L. K., Parfrey, L. W. and Knight, R. (2012). “The impact of the gut microbiota on human health: an integrative view.” Cell 148(6): 1258–1270 DOI: https://doi-org.utk.idm.oclc.org/10.1016/j.cell.2012.01.035.

11. Do, T. T., Delaney, S. and Walsh, F. (2019). “16S rRNA gene based bacterial community structure of wastewater treatment plant effluents.” FEMS microbiology letters 366(3) DOI: https://10.1093/femsle/fnz017.

12. Fierer, N., Holland-Moritz, H., Alexiev, A., Batther, H., Dragone, N. B., Friar, L., Gebert, M. J., Gering, S., Henley, J. B. and Jech, S. (2022). “A metagenomic investigation of spatial and temporal changes in sewage microbiomes across a university campus.” Msystems 7(5): e00651–00622 DOI: https://doi-org.utk.idm.oclc.org/10.1128/msystems.00651-22.

13. Fujii, M., Kojima, H., Iwata, T., Urabe, J. and Fukui, M. (2012). “Dissolved Organic Carbon as Major Environmental Factor Affecting Bacterioplankton Communities in Mountain Lakes of Eastern Japan.” Microbial ecology 63(3): 496–508 DOI: 10.1007/s00248-011-9983-8.

14. Furet, J.-P., Firmesse, O., Gourmelon, M., Bridonneau, C., Tap, J., Mondot, S., Doré, J. and Corthier, G. (2009). “Comparative assessment of human and farm animal faecal microbiota using real-time quantitative PCR.” FEMS microbiology ecology 68(3): 351–362 DOI: 10.1111/j.1574-6941.2009.00671.x.

15. Gallardo-Escárate, C., Valenzuela-Muñoz, V., Núñez-Acuña, G., Valenzuela-Miranda, D., Benaventel, B. P., Sáez-Vera, C., Urrutia, H., Novoa, B., Figueras, A., Roberts, S., Assmann, P. and Bravo, M. (2021). “The wastewater microbiome: A novel insight for COVID-19 surveillance.” Science of the Total Environment 764: 142867 DOI: 10.1016/j.scitotenv.2020.142867.

16. Gu, S., Chen, Y., Wu, Z., Chen, Y., Gao, H., Lv, L., Guo, F., Zhang, X., Luo, R. and Huang, C. (2020). “Alterations of the gut microbiota in patients with coronavirus disease 2019 or H1N1 influenza.” Clinical Infectious Diseases 71(10): 2669–2678 DOI: https://doi-org.utk.idm.oclc.org/10.1093/cid/ciaa709.

17. Huttenhower, C., Gevers, D., Knight, R., Abubucker, S., Badger, J. H., Chinwalla, A. T., Creasy, H. H., Earl, A. M., FitzGerald, M. G., Fulton, R. S., Giglio, M. G., Hallsworth-Pepin, K., Lobos, E. A., Madupu, R., Magrini, V., Martin, J. C., Mitreva, M., Muzny, D. M., Sodergren, E. J., Versalovic, J., Wollam, A. M., Worley, K. C., Wortman, J. R., Young, S. K., Zeng, Q., Aagaard, K. M., Abolude, O. O., Allen-Vercoe, E., Alm, E. J., Alvarado, L., Andersen, G. L., Anderson, S., Appelbaum, E., Arachchi, H. M., Armitage, G., Arze, C. A., Ayvaz, T., Baker, C. C., Begg, L., Belachew, T., Bhonagiri, V., Bihan, M., Blaser, M. J., Bloom, T., Bonazzi, V., Paul Brooks, J., Buck, G. A., Buhay, C. J., Busam, D. A., Campbell, J. L., Canon, S. R., Cantarel, B. L., Chain, P. S. G., Chen, I. M. A., Chen, L., Chhibba, S., Chu, K., Ciulla, D. M., Clemente, J. C., Clifton, S. W., Conlan, S., Crabtree, J., Cutting, M. A., Davidovics, N. J., Davis, C. C., DeSantis, T. Z., Deal, C., Delehaunty, K. D., Dewhirst, F. E., Deych, E., Ding, Y., Dooling, D. J., Dugan, S. P., Michael Dunne, W., Scott Durkin, A., Edgar, R. C., Erlich, R. L., Farmer, C. N., Farrell, R. M., Faust, K., Feldgarden, M., Felix, V. M., Fisher, S., Fodor, A. A., Forney, L. J., Foster, L., Di Francesco, V., Friedman, J., Friedrich, D. C., Fronick, C. C., Fulton, L. L., Gao, H., Garcia, N., Giannoukos, G., Giblin, C., Giovanni, M. Y., Goldberg, J. M., Goll, J., Gonzalez, A., Griggs, A., Gujja, S., Kinder Haake, S., Haas, B. J., Hamilton, H. A., Harris, E. L., Hepburn, T. A., Herter, B., Hoffmann, D. E., Holder, M. E., Howarth, C., Huang, K. H., Huse, S. M., Izard, J., Jansson, J. K., Jiang, H., Jordan, C., Joshi, V., Katancik, J. A., Keitel, W. A., Kelley, S. T., Kells, C., King, N. B., Knights, D., Kong, H. H., Koren, O., Koren, S., Kota, K. C., Kovar, C. L., Kyrpides, N. C., La Rosa, P. S., Lee, S. L., Lemon, K. P., Lennon, N., Lewis, C. M., Lewis, L., Ley, R. E., Li, K., Liolios, K., Liu, B., Liu, Y., Lo, C.-C., Lozupone, C. A., Dwayne Lunsford, R., Madden, T., Mahurkar, A. A., Mannon, P. J., Mardis, E. R., Markowitz, V. M., Mavromatis, K., McCorrison, J. M., McDonald, D., McEwen, J., McGuire, A. L., McInnes, P., Mehta, T., Mihindukulasuriya, K. A., Miller, J. R., Minx, P. J., Newsham, I., Nusbaum, C., O’Laughlin, M., Orvis, J., Pagani, I., Palaniappan, K., Patel, S. M., Pearson, M., Peterson, J., Podar, M., Pohl, C., Pollard, K. S., Pop, M., Priest, M. E., Proctor, L. M., Qin, X., Raes, J., Ravel, J., Reid, J. G., Rho, M., Rhodes, R., Riehle, K. P., Rivera, M. C., Rodriguez-Mueller, B., Rogers, Y.-H., Ross, M. C., Russ, C., Sanka, R. K., Sankar, P., Fah Sathirapongsasuti, J., Schloss, J. A., Schloss, P. D., Schmidt, T. M., Scholz, M., Schriml, L., Schubert, A. M., Segata, N., Segre, J. A., Shannon, W. D., Sharp, R. R., Sharpton, T. J., Shenoy, N., Sheth, N. U., Simone, G. A., Singh, I., Smillie, C. S., Sobel, J. D., Sommer, D. D., Spicer, P., Sutton, G. G., Sykes, S. M., Tabbaa, D. G., Thiagarajan, M., Tomlinson, C. M., Torralba, M., Treangen, T. J., Truty, R. M., Vishnivetskaya, T. A., Walker, J., Wang, L., Wang, Z., Ward, D. V., Warren, W., Watson, M. A., Wellington, C., Wetterstrand, K. A., White, J. R., Wilczek-Boney, K., Wu, Y., Wylie, K. M., Wylie, T., Yandava, C., Ye, L., Ye, Y., Yooseph, S., Youmans, B. P., Zhang, L., Zhou, Y., Zhu, Y., Zoloth, L., Zucker, J. D., Birren, B. W., Gibbs, R. A., Highlander, S. K., Methé, B. A., Nelson, K. E., Petrosino, J. F., Weinstock, G. M., Wilson, R. K., White, O. and The Human Microbiome Project, C. (2012). “Structure, function and diversity of the healthy human microbiome.” Nature 486(7402): 207-214 DOI: 10.1038/nature11234.

18. Li, Y., Ash, K. T., Joyner, D. C., Williams, D. E., Alamilla, I., McKay, P., Iler, C., Green, B., Kara-Murdoch, F. and Swift, C. M. (2023). “Decay of enveloped SARS-CoV-2 and non-enveloped PMMoV RNA in raw sewage from university dormitories.” Frontiers in Microbiology 14: 1144026 DOI: 10.3389/fmicb.2023.1144026.

19. Lindström, E. S., Agterveld, M. P. K.-V. and Zwart, G. (2005). “Distribution of Typical Freshwater Bacterial Groups Is Associated with pH, Temperature, and Lake Water Retention Time.” Applied and Environmental Microbiology 71(12): 8201–8206 DOI: 10.1128/AEM.71.12.8201-8206.2005.

20. Liu, Q., Mak, J. W. Y., Su, Q., Yeoh, Y. K., Lui, G. C.-Y., Ng, S. S. S., Zhang, F., Li, A. Y. L., Lu, W., Hui, D. S.-C., Chan, P. K., Chan, F. K. L. and Ng, S. C. (2022). “Gut microbiota dynamics in a prospective cohort of patients with post-acute COVID-19 syndrome.” Gut 71(3): 544–552 DOI: 10.1136/gutjnl-2021-325989.

21. Newton, R. J., McLellan, S. L., Dila, D. K., Vineis, J. H., Morrison, H. G., Eren, A. M. and Sogin, M. L. (2015). “Sewage reflects the microbiomes of human populations.” mBio 6(2): 10.1128/mbio.02574-02514.

22. Numberger, D., Ganzert, L., Zoccarato, L., Mühldorfer, K., Sauer, S., Grossart, H.-P. and Greenwood, A. D. (2019). “Characterization of bacterial communities in wastewater with enhanced taxonomic resolution by full-length 16S rRNA sequencing.” Scientific Reports 9(1): 9673 DOI: 10.1038/s41598-019-46015-z.

23. Oluseyi Osunmakinde, C., Selvarajan, R., Mamba, B. B. and Msagati, T. A. M. (2019). “Profiling Bacterial Diversity and Potential Pathogens in Wastewater Treatment Plants Using High-Throughput Sequencing Analysis.” Microorganisms 7(11): 506 DOI: https://doi-org.utk.idm.oclc.org/10.3390/microorganisms7110506.

24. Poopedi, E., Singh, T. and Gomba, A. (2023). “Potential Exposure to Respiratory and Enteric Bacterial Pathogens among Wastewater Treatment Plant Workers, South Africa.” International Journal of Environmental Research and Public Health 20(5): 4338 DOI: https://doi-org.utk.idm.oclc.org/10.3390/ijerph20054338.

25. Turnbaugh, P. J., Ley, R. E., Mahowald, M. A., Magrini, V., Mardis, E. R. and Gordon, J. I. (2006). “An obesity-associated gut microbiome with increased capacity for energy harvest.” Nature 444(7122): 1027–1031 DOI: https://10.1038/nature05414.

26. Yeoh, Y. K., Zuo, T., Lui, G. C.-Y., Zhang, F., Liu, Q., Li, A. Y., Chung, A. C., Cheung, C. P., Tso, E. Y., Fung, K. S., Chan, V., Ling, L., Joynt, G., Hui, D. S.-C., Chow, K. M., Ng, S. S. S., Li, T. C.-M., Ng, R. W., Yip, T. C., Wong, G. L.-H., Chan, F. K., Wong, C. K., Chan, P. K. and Ng, S. C. (2021). “Gut microbiota composition reflects disease severity and dysfunctional immune responses in patients with COVID-19.” Gut 70(4): 698–706 DOI: 10.1136/gutjnl-2020-323020.

27. Zhang, F., Lau, R. I., Liu, Q., Su, Q., Chan, F. K. L. and Ng, S. C. (2023). “Gut microbiota in COVID-19: key microbial changes, potential mechanisms and clinical applications.” Nature Reviews Gastroenterology & Hepatology 20(5): 323–337 DOI: 10.1038/s41575-022-00698-4.

28. Zuo, T., Zhang, F., Lui, G. C., Yeoh, Y. K., Li, A. Y., Zhan, H., Wan, Y., Chung, A. C., Cheung, C. P. and Chen, N. (2020). “Alterations in gut microbiota of patients with COVID-19 during time of hospitalization.” Gastroenterology 159(3): 944–955. e948 DOI: 10.1053/j.gastro.2020.05.048.

