## Supplementary figures and images for "SARS-CoV-2 virus in Raw Wastewater from Student Residence Halls with concomitant 16S rRNA Bacterial Community Structure changes"

### Supplemental Figure S1

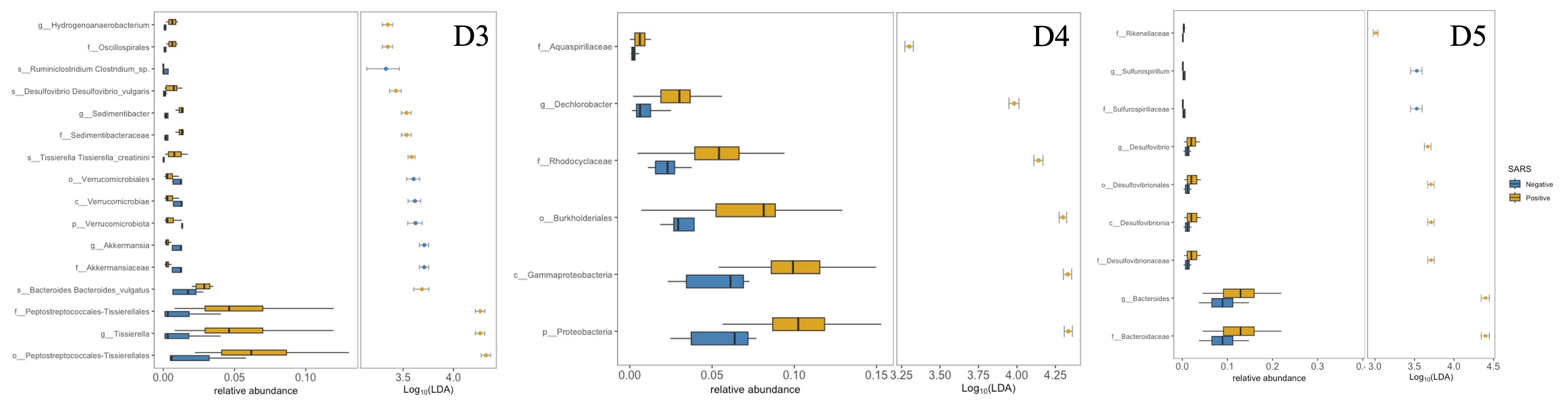
